# Indirect impact on number of admitted children with COVID-19 in the first three months of emergency use of two vaccines, in Rio de Janeiro, Brazil

**DOI:** 10.1101/2021.07.01.21259330

**Authors:** CGB Fonseca, ALTG Costa, MDM Esteves, BRR de Carvalho, CV Souza, CH Teixeira, AR Araujo da Silva

**Affiliations:** Universidade Federal Fluminense; Infection Control Committee- Prontobaby-Group

**Keywords:** COVID 19, pediatrics, clinical manifestations, hospitalization, vaccine

## Abstract

**Introduction:** Since January 19, 2021, two vaccines against SARS-COV-2 are available in Brazil for emergency use for selected groups, not including children.

**Aim:** To describe indirect impact on the number of hospitalized children with COVID-19 in the first three months after beginning of emergency use of two vaccines.

**Methods:** A retrospective study was conducted in children (0-18 years), admitted in two pediatric hospitals of Rio de Janeiro city, between January and April 2021 with confirmed COVID-19 by reverse transcription polymerase chain reaction or serological tests. Number of cases, clinical signs, symptoms and outcomes were compared with the first wave of disease (April-June 2020). A p value of less than .05 were considered was significant.

**Results:** The number of total admitted patients (with all diseases) were 1097 in 2020 period, being 46 (4.2%) of them with confirmed COVID-19, and 2187 in the 2021 period, with 47 (2.1%) cases (p=0.006). Predominant respiratory symptoms were present in 29/46 (63%) as the main presentation in 2020 patients and 37/47 (78.7%) in 2021 children (p=0.09). The main symptoms were fever, cough and dyspnoea in both periods. The median of lenght of stay after diagnosis were 4 days in 2020 and 2021 (p=0.9). Just one patient died in 2021.

**Conclusion:** There was reduction of relative percentage of admitted confirmed pediatric cases in the the first three months of emergency use of two vaccines against SARS-COV-2, but it’s uncertain to atribute this finding to vaccination due to high circulation of the virus in the city.

## Introduction

Until June 18, 2021 more than 177 million of people were diagnosed with COVID-19 in all countries, with at least 3,8 million of deaths. ^1^ The pandemic of SARS-COV-2 stills in progress in May 2021, despite global efforts to reduce impact of disease around the world. ^1^

Several non-pharmaceutical interventions (NPIs) were implemented wordwide to mitigate the spread of SARS-CoV-2, considering absence of vaccine or effective antiviral in the first year of pandemics and includes closure of educational institutions, national lockdown, mass gathering cancellation, border restriction, quarantine and others. ^2,3^

After massive research simultaneously conducted in different countries, companies and universities, the first vaccine (Pfizer–BioNTech) for COVID-19 prevention was approved in early December 2020, by United Kingdom’s Medicines and Healthcare products Regulatory Agency (MHRA) for selected groups. ^4^ Since then, other vaccines were approved for different countries including Brazil, but effects of reducing cases needs to be studied, including subgroups not included in vaccination as children.

Considering the use of two vaccines for COVID-19 prevention since January 2021, in Brazil, our aim is to describe indirect impact on the number of hospitalized children with COVID-19 in the first three months after beginning of emergency use of two vaccines.

## Methods

### Study design and setting

We conducted a retrospective cohort study in two pediatric hospitals in Rio de Janeiro, city, Brazil. Both hospitals are private units destined exclusively to pediatric patients.

Unit 1 is a 135-bed hospital located at North zone of the city, that receives clinical and surgical patients referred from its own emergency and from other services. Unit 2 is a 39-bed hospital, located at South zone of the city, with the same profile of unit 1.

### Period of the study

Two periods were analysed: the first one comprised the first day of Brazilian epidemiological week 15 (April 5, 2020) and the last day of epidemiological week 27 (July 4, 2020). This period corresponds to the first wave of COVID-19, according to the historical series of confirmed cases attending in both hospitals. The second period comprises the first day of epidemiological week 3 of 2021 (January 17^th^) and last day of epidemiological week 15 of the same year (April 17^th^). This period corresponds to the first three months after beginning of vaccination in Rio de Janeiro city.

In both periods studied, higher number of COVID-19 cases were detected in the city. ^5^

### Inclusion and exclusion criteria

All patients admitted with COVID-19 in both hospitals were included in analysis. Children admitted but transferred to the other hospitals within the first 24h were excluded.

### COVID-19 vaccines available and priority groups

Two vaccines were available by emergency use in Brazil, by The Brazilian Health Regulatory Agency (ANVISA): CoronaVac ® and Oxford/AstraZeneca ® and one for definitive use (Pfizer-BioNTech ®), at the end of April 2021. ^6^ The first doses of vaccines were administered in Rio de Janeiro, in January 20, 2021, but until the end of April 2021, Pfizer-BioNTech vaccine was not available for population and not included in the current analysis

Until April 14 2021, the following groups received at least one dose of vaccine: persons older than 63 years and healthcare workers (all ages) with direct contact with patients. No vaccine was available for children

### Diagnostic of COVID-19 infection

A patient was considered as a confirmed case of COVID-19 in the presence of positive polymerase chain reaction (PCR), positive IgM serology or positive antigen reaction. For epidemiological analyses, we considered date of beginning of the symptoms.

### Data source and statistical analysis

The two periods were analysed regarding the following variables: Total number of admitted patients (with all diseases), total number of admitted patients with confirmed COVID-19, the main clinical symptoms present on admission and number of deaths

A descriptive analysis was performed using Microsoft Excel. When appropriate, we used Chi square test for categorical variables and Mann–Whitney U test for continuous variables. A value of p less than .05 were considered as statistical significant.

### Ethical aspects

The study was submitted and approved by Ethics Committee of Faculty of Medicine (Universidade Federal Fluminense) and Prontobaby Group, under number 4.100.232 dated from June 20, 2020.

## Results

In the first period analysed (April 5,2020-July 4, 2020), 1097 children were admitted, being 46 (4.2%) with confirmed COVID-19 infection. Considering the second of period of analysis (January 17, 2021-April 17, 2021), 2187 children were admitted, with 47 (2.1%) confirmed COVID-19 infection (p=0.0006). Demographic data about presentation of symptoms and outcomes are presented in table 1

**Table 1-.** Demographic data of pediatric patients admitted with COVID-19 in the first wave and first 12 weeks after beginning of emergency vaccine in Rio de Janeiro, city

|  | Confimed cases first<br>wave<br>N= 46 | Confirmed cases of first<br>12 weeks after beginning<br>of emergency vaccination<br>N=47 | P<br>value |
| --- | --- | --- | --- |
| Male gender (%) | 28 (60.8) | 25 (53.2) | 0.46 |
| Median age ( in months) | 86.5 | 41 | 0.12 |
| **Clinical presentation** |  |  |  |
| Predominant respiratory symptoms (%) | 29 (63) | 37 (78.7) | 0.09 |
| Predominant non-respiratory symptoms (%) | 17 (37) | 10 (21.2) |  |
| **Outcomes** |  |  |  |
| Discharged | 46 (100) | 46 ( 97.9) | 0.32 |
| Dead | 0 | 1 ( 2.1) |  |
| **Lenght of stay after diagnosis** (median in days) | 4 | 4 | 0.90 |

The main symptoms in the first wave patients were fever in 36/46 (78.3%) of patients, dyspnoea in 24/46 (52.2%) and cough in 23/46 (50%). In the second period of analysis the most common symptoms were fever in 35/47 (74.5%) children, cough in 34/47 (72.3%) and dyspnoea in 25/47 (53.2%) patients

The graph one shows the cases since the first one diagnosed in 2020 until April 17, 2021, considering epidemiological weeks of Brazil surveillance

**Graph 1.**
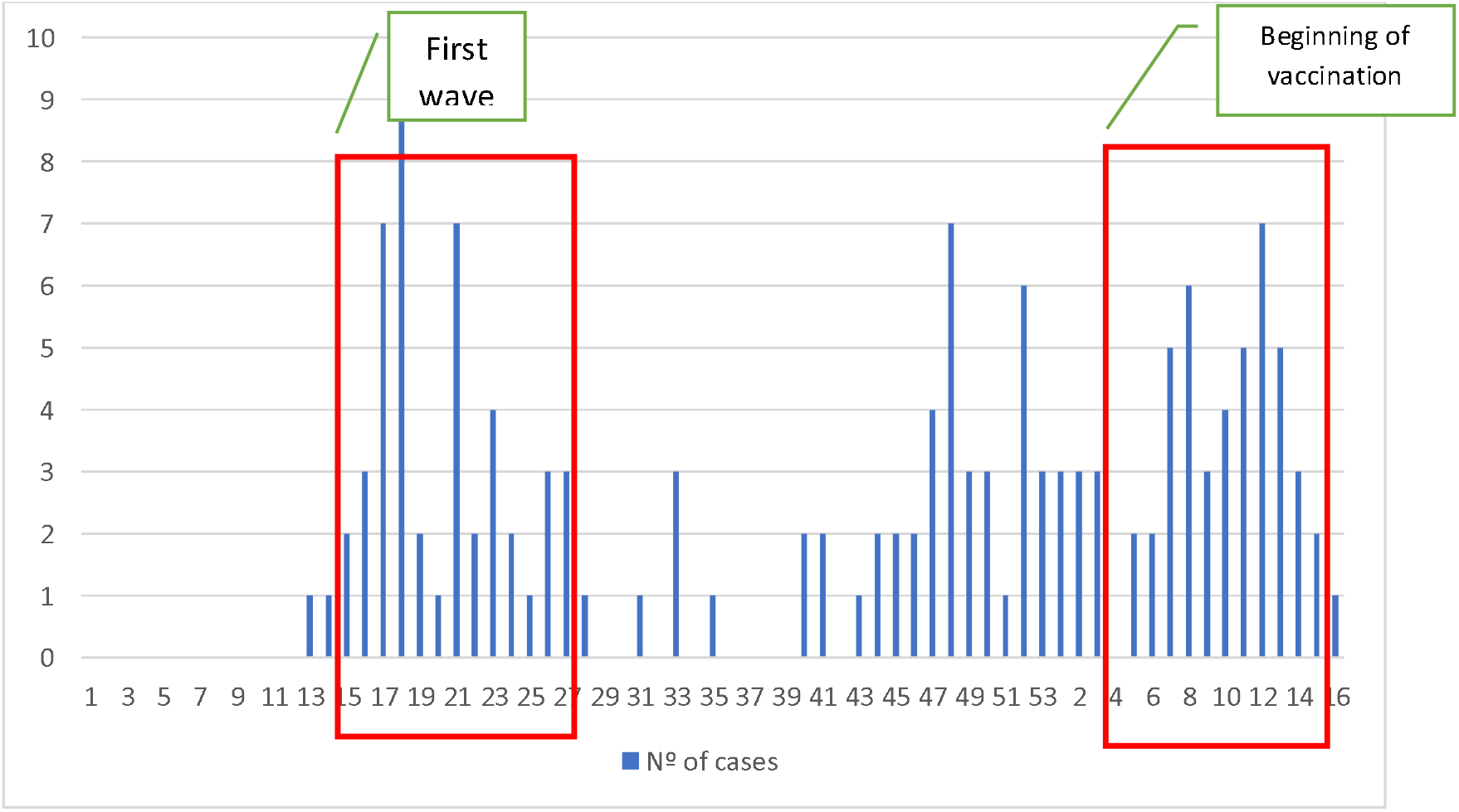
Number of admitted pediatric confirmed COVID cases considering epidemiological weeks of Brazil surveillance (2020-2021)

## Discussion

According to the last Brazilian Epidemiological Bulletin relative to COVID data, children represent around 1.4% of acute respiratory distress syndrome (ARDS) cases and 0.35 % of deaths atributted to COVID ARDS, respectively during 2021 year in Brazil. But, despite of relative few number of cases and deaths, the pandemic still uncontrolled in the whole country. ^7^

Starting in end of 2020, systematic vaccination of selected groups is increasing in the world as a strategy to limit the spread of SARS-COV-2 in addition to NPIs. ^4^ Brazil began his vaccination program in January 2021, for elderly people and healthcare personnel and it’s possible that some indirect impact about reduction of pediatric cases could be verified in children, due to family composition in Brazil, where grandparents participate of children care. ^8^

Considering the two periods of analysis in our study, number of admitted children due to the all causes increased in 2021 compared with previous year. We could atribute this finding to the restriction of circulation in Rio de Janeiro in the 2020 year and flexibilization of coletive rules in the second period. Other countries as Singapore and Canada also experienced reduction of visities to emergency department and hospitalizations. ^9,10^

COVID-19 is a mild/severe infection in children and we verified that the main symptoms of disease (fever, cough and dyspnoea) were similar in both years studied, confirming that disease is a predominant illness of respiratory tract, with some cases of unsual presentation as multisystem inflammatory syndrome (MIS-C) and just one death. A recent meta-analysis about COVID-19 concluded that children and/or adolescents tend to have a mild COVID-19 course with a good prognosis when compared to adults.^11^

The absolute number of COVID-19 cases were similar in both periods analysed, but when we compared relative percentage of cases, considering total number of admitted patients, there was a statistically significant reduction of confirmed cases.

Vaccination against SARS-COV-2 is in different stages around the world and countries that have been started his programm earlier, covering higher percentage of population, obtained promising results. One of the first reports about effects of vaccination verified a larger and earlier decrease in COVID-19 cases and hospitalization in individuals older than 60 years, followed by younger age groups. ^12^ In United Kingdom, a model suggests substantial reductions in hospital and intensive care units admissions would not occur until three/four months after beginning of COVID-19 vaccination in that country. ^13^

The positive impact about relative percentage of admitted children during the study period could be na initial marker of vaccination effects, but natural decrease of number of cases, considering a possible seasonality of the vírus, could be not foreclosed.

Our study has some limitations. The first one is that our data showed pattern of admissions in two pediatric hospital of the city. But considering that both hospitals are the biggest institutions dedicated exclusively to the children care in the city, that data could reflect the true dynamics of SARS-COV-2 circulation in the city. The second limitation is possible influence of knowledge acquistion of pediatricians to treat COVID-19, during the first year of pandemic, resulting in fewer relative percentage of admitted children with COVID-19 in 2021. Despite this possibility could be not rulled out, until the current date, no drug was approved in Brazil to avoid or reduce pediatric COVID-19 admissions.

## Conclusion

There was reduction of relative percentage of admitted confirmed pediatric cases in the the first three months of emergency use of two vaccines against SARS-COV-2, but it’s uncertain to atribute this finding to vaccination due to high circulation of the virus in the city.

## Data Availability

All data are available for further analysis

## Acknowlegdment

We thank to Mario Eduardo Viana and Luisa Benigno Barbosa Araujo da Silva for supporting this research.

